# Right caudate deactivation during reward anticipation predicts elevated risk for psychosis in adolescents

**DOI:** 10.1101/2024.10.02.24314810

**Authors:** Pritha Sen, Franziska Knolle

**Author notes:** Corresponding author: Dr. Franziska Knolle, Department of Diagnostic and Interventional Neuroradiology, School of Medicine and Health, Klinikum rechts der Isar, Technical University Munich, 81675 Munich, Germany.

## Abstract

**Background:** Schizophrenia is associated with abnormalities in neurodevelopmental processes. Furthermore, dysfunctional neural circuits involved in reward processing may be linked to the development of symptoms in schizophrenia and are predictive of long-term functional outcome. It is however unknown whether neural signatures of reward processing are detectable in children with an increased risk of psychosis and/or predictive of their prodromal psychosis score.

**Methods:** Using data from the ABCD study 4.1, we defined a healthy control (N=50) and a risk for psychosis (N=50) group with a Prodromal Psychosis Syndrome (PPS) score>3 at baseline (9– 10 years) and 2^nd^-year-follow-up (11–12 years). While undergoing functional MR-imaging, all children completed the Monetary Incentive Delay task. Using the preprocessed ABCD-data, we explore whether behaviour and brain activations for reward and loss anticipation in areas underlying reward processing differed between groups and time-points. Furthermore, we investigated whether those brain activations that showed differences between the groups were predictive of later PPS scores.

**Results:** While behavioural results did not differ, we found that at-risk children demonstrated lower activation during reward anticipation in the caudate for both timepoints, and the nucleus accumbens, the putamen, the dorsolateral and the ventral medial prefrontal cortex for the 2^nd^-year-follow-up compared to controls. Regression analysis revealed that right caudate deactivation for both timepoints was predictive of later PPS scores.

**Conclusion:** This study reveals that neural alterations during reward anticipation are detectable in children at-risk of psychosis. These dysfunctions in neural activation patterns may serve as a potential predictive biomarker for psychosis.

## Introduction

Schizophrenia is one of the leading causes of years lived with disabilities [1] causing significant economic burden to the society [1], and reducing life expectancy of individuals by approximately 15 years [1–3]. While patients with a longer duration of untreated psychosis tend to have poorer prognosis (Marshall et al., 2005), individuals with early detection of psychosis show better treatment responses (Li et al., 2022) and significantly reduced conversion rates to frank psychosis [4,5]. Thus, current research goals and strategies aim at identifying predictive markers for early identification and treatment for individuals at-risk of developing psychosis [6–8], which has hence been regarded as the most promising strategy to ameliorate the devastating consequences of psychotic disorders.

Although hallucinations and delusions are the primary diagnostic features of psychotic disorders such as schizophrenia, impairments in reward-learning are better predictors of functional outcomes [9,10]. Reward-learning is essential for everyday life as it underlies almost all of our choices. It involves neural feedback and feedforward systems critical for prediction, motivation, goal-directed behaviour, as well as overall cognition [11]. Generally, it has been shown that alterations in the brain’s reward system may contribute to the symptoms of schizophrenia [12–14]. Studies on adults at different stages of psychosis and schizophrenia have revealed behavioural alterations (e.g. [11,15–17]) and dysfunctions in the cortico-subcortical circuits underlying reward processing (e.g. [18–21]).

In schizophrenia patients, reduced activations in response to reward anticipations are reported consistently in the ventral striatum [22–25], along with hypoactivation in the insula, amygdala, anterior cingulate cortex (ACC) and dorsolateral prefrontal cortex (dlPFC) [12,24] and with hypoactivation in the medial cingulate cortex, precentral gyrus, and superior temporal [25]. Also, in earlier stages of psychosis dysfunctional reward anticipation processing has been reported. In individuals with subclinical psychotic experiences, Michielse and colleagues [26] showed reduced activations during reward anticipation in the right insula, putamen and supramarginal gyrus. Similarly, reduced reward anticipation activation in the dlPFC and the ventral striatum have been reported in antipsychotic naïve first episode psychosis patients [27,28]. Reduced striatal activations were found to be associated with the severity of positive symptoms early psychosis patients, suggesting that these abnormalities are present from the beginning of the disease [27].

There is, however, very little evidence whether similar processing alterations can already be detected in children with increased risk of psychosis. One study [29] explored whether psychotic-like experiences (PLEs) and PLE-related distress in 6718 children aged 9-10 from the ABCD-cohort were associated with reward-related neural alterations in the nucleus accumbens during the completion of an imaging version of the Monetary Incentive Delay (MID) task [30]. Their analysis did not reveal any significant associations [29], which may be due to the low endorsement of PLEs across this large sample with a disproportional high number of those without any PLEs. We therefore set out to explore this question with a different approach. We defined two strictly matched groups of 50 individuals each, one healthy group without any endorsement of PPLs and no psychiatric history and a group at-risk for psychosis with PPLs, measured with the Prodromal Psychosis Syndrome (PPS) scores greater than 3 at baseline (9–10 years) and 2^nd^-year-follow-up (11–12 years). We then investigated whether reward or loss anticipation was different across the two groups in regions crucial to reward-learning (i.e., bilateral caudate, nucleus accumbens, thalamus, putamen, dlPFC and vmPFC). We hypothesized that the at-risk group would show hypoactivation in the prefrontal cortical and striatal regions, especially in the nucleus accumbens, during reward anticipation in the MID task. Furthermore, we expected that potential neural alterations were predictive of later PLEs.

## Methods

### Participants

Participants were included from the ongoing large-scale longitudinal study, Adolescent Brain and Cognitive Development (ABCD) (release 4.0; https://abcdstudy.org/) (11,876 subjects). Currently, complete datasets of four time-points have been released, with imaging data available at baseline and the 2^nd^ year follow-up. A subset from the baseline and 2^nd^ year follow-up imaging and behavioural data for MID task was included in the current study.

We selected two strictly matched groups, a healthy control group and at-risk for psychosis group. Risk of developing psychosis was examined using the PPS score which is the sum of the number of “yes” responses to the questions in the Prodromal Questionnaire-Brief Child version (PQ-BC) as a measure of self-reported PLEs. For each “yes”-response, a distress score was assessed asking how much the experience bothered them on a scale of 1-5. The total distress score is calculated as the total number of endorsed questions weighted by the level of distress. Research suggests that individuals with PPS > 3 or distress score > 6 are at an increased risk of developing psychotic disorders [31,32]. Therefore, our risk group was defined by selecting 50 individuals with PPS score > 3 in both baseline and 2^nd^ year follow-up time-points (Figure 1). The distress scores for all time-points were also calculated to ensure that the risk individuals lay above the cut-off. Clinical and demographic scores are presented in Table 1.

**Figure 1.**
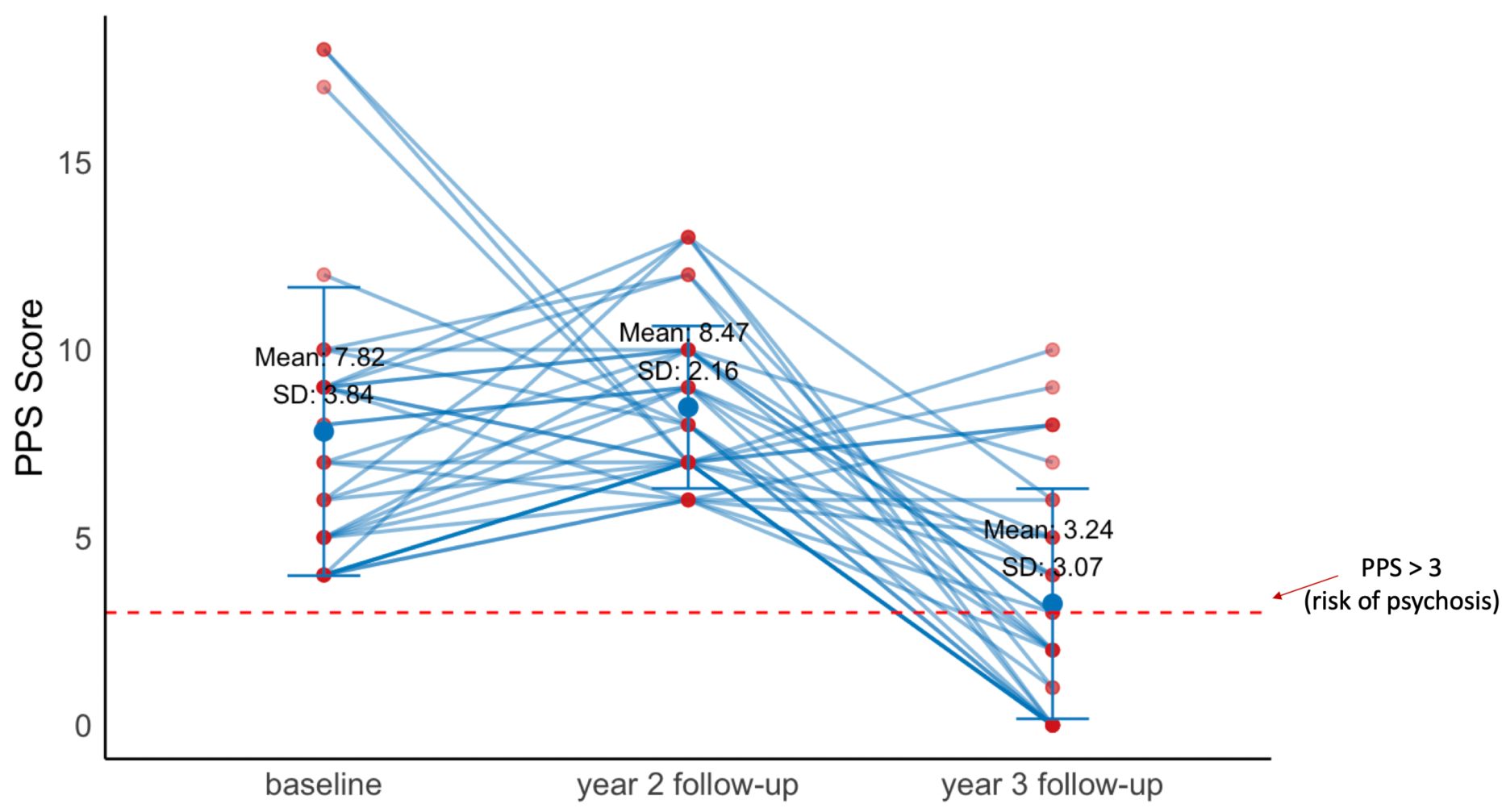
Progression in PPS scores across for the at-risk group for the timepoints investigated in this study. The error bar represents mean and standard deviation. The red dotted represents PPS = 3, based on Karcher et al, 2023, PPS scores > 3 as an early risk for psychosis [31].

**Table 1.**
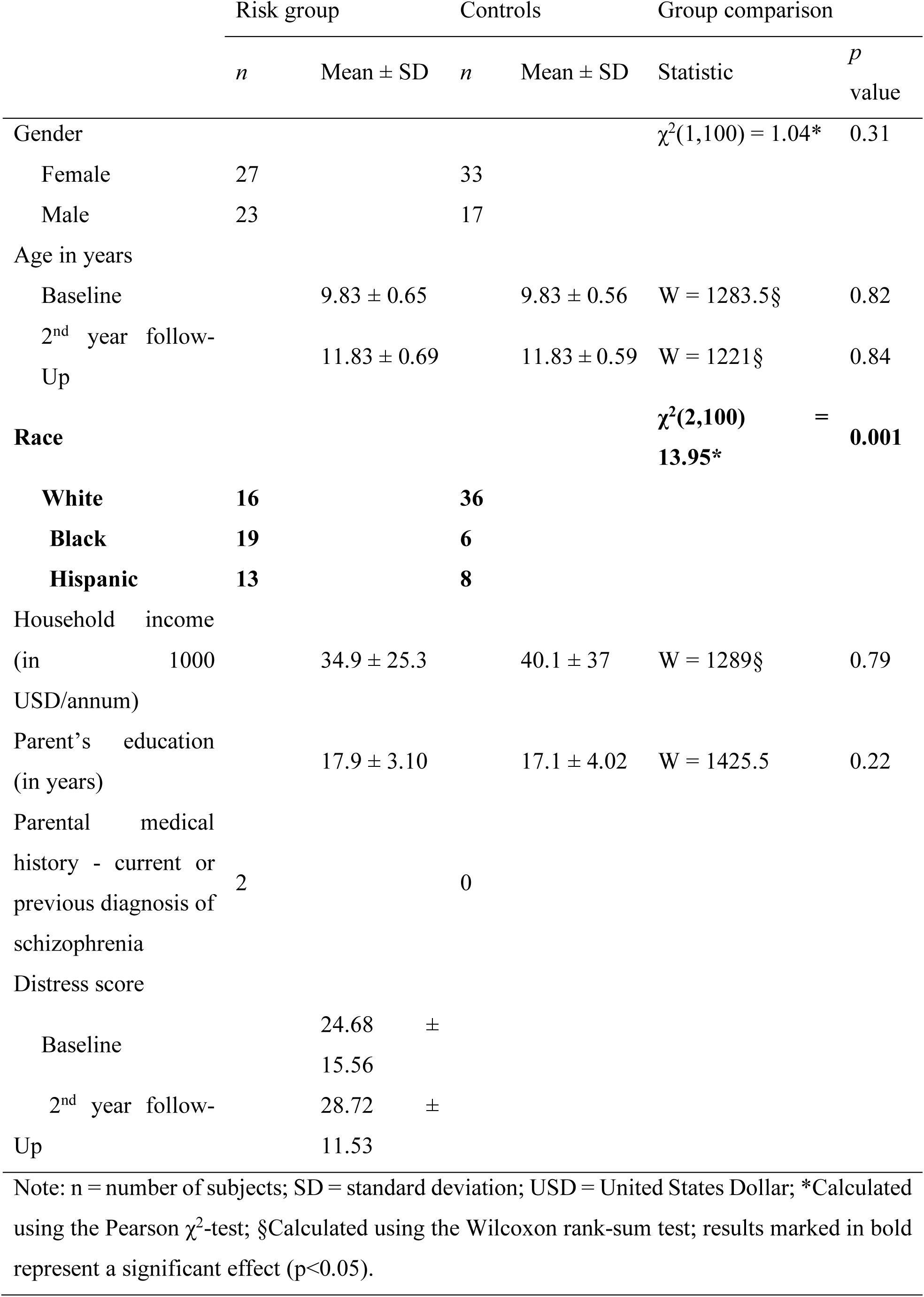
Demographic information for risk and control subjects.

|  | Risk group |  | Controls |  | Group comparison |  |
| --- | --- | --- | --- | --- | --- | --- |
| | *n* | Mean ± SD | *n* | Mean ± SD | Statistic | *p* value |
| Gender | | | | | $\chi^2(1,100) = 1.04^*$ | 0.31 |
| Female | 27 |  | 33 |  |  |  |
| Male | 23 |  | 17 |  |  |  |
| Age in years |  |  |  |  |  |  |
| Baseline | | 9.83 ± 0.65 | | 9.83 ± 0.56 | W = 1283.5§ | 0.82 |
| 2<sup>nd</sup> year follow-Up | | 11.83 ± 0.69 | | 11.83 ± 0.59 | W = 1221§ | 0.84 |
| Race | | | | | $\chi^2(2,100) = 13.95^*$ | **0.001** |
| White | **16** |  | **36** |  |  |  |
| Black | **19** |  | **6** |  |  |  |
| Hispanic | **13** |  | **8** |  |  |  |
| Household income<br>(in 1000 USD/annum) | | 34.9 ± 25.3 | | 40.1 ± 37 | W = 1289§ | 0.79 |
| Parent's education<br>(in years) | | 17.9 ± 3.10 | | 17.1 ± 4.02 | W = 1425.5 | 0.22 |
| Parental medical<br>history - current or<br>previous diagnosis of<br>schizophrenia | 2 |  | 0 |  |  |  |
| Distress score |  |  |  |  |  |  |
| Baseline | | 24.68 ± 15.56 | | | | |
| 2<sup>nd</sup> year follow-Up | | 28.72 ± 11.53 | | | | |
| Note: n = number of subjects; SD = standard deviation; USD = United States Dollar; *Calculated using the Pearson $\chi^2$-test; §Calculated using the Wilcoxon rank-sum test; results marked in bold represent a significant effect ($p < 0.05$). | | | | | | |

For the control group, individuals with PPS = 0 at all four time-points were selected, from which participants whose parents had a history of mental illnesses were excluded using the ABCD Parent Diagnostic Interview for DSM-5 Full. Finally, a random sample of 50 individuals was selected as the control group, matched for gender, age, household income and parental education to the risk group. The two group, however, differed in ethnicity. Matching both groups was not possible as too few individuals fulfilled all criteria. Demographic information for both the groups is displayed in Table 1.

### Task description

In the MID task (Figure 2), participants were required to earn monetary rewards through rapid motor responses while completing 100 trials. Before the task began, the participants learnt the association between shapes, which are presented as cues, and large ($5, -$5), small ($0.20, - $0.20), or neutral ($0) rewards or losses. Each trial began with a cue stimulus (2000ms), predicting either large or small rewards or losses. This was followed by a jittered delay (1500-4000ms), representing the anticipation phase. Then, a variable target was presented for 150-500ms, and participants were required to respond as quickly as possible by clicking a button to either win money or avoid losing it. Finally, feedback was provided on the outcome. Details of this task are described elsewhere [33]. Participants performed this task inside an fMRI scanner.

**Figure 2.**
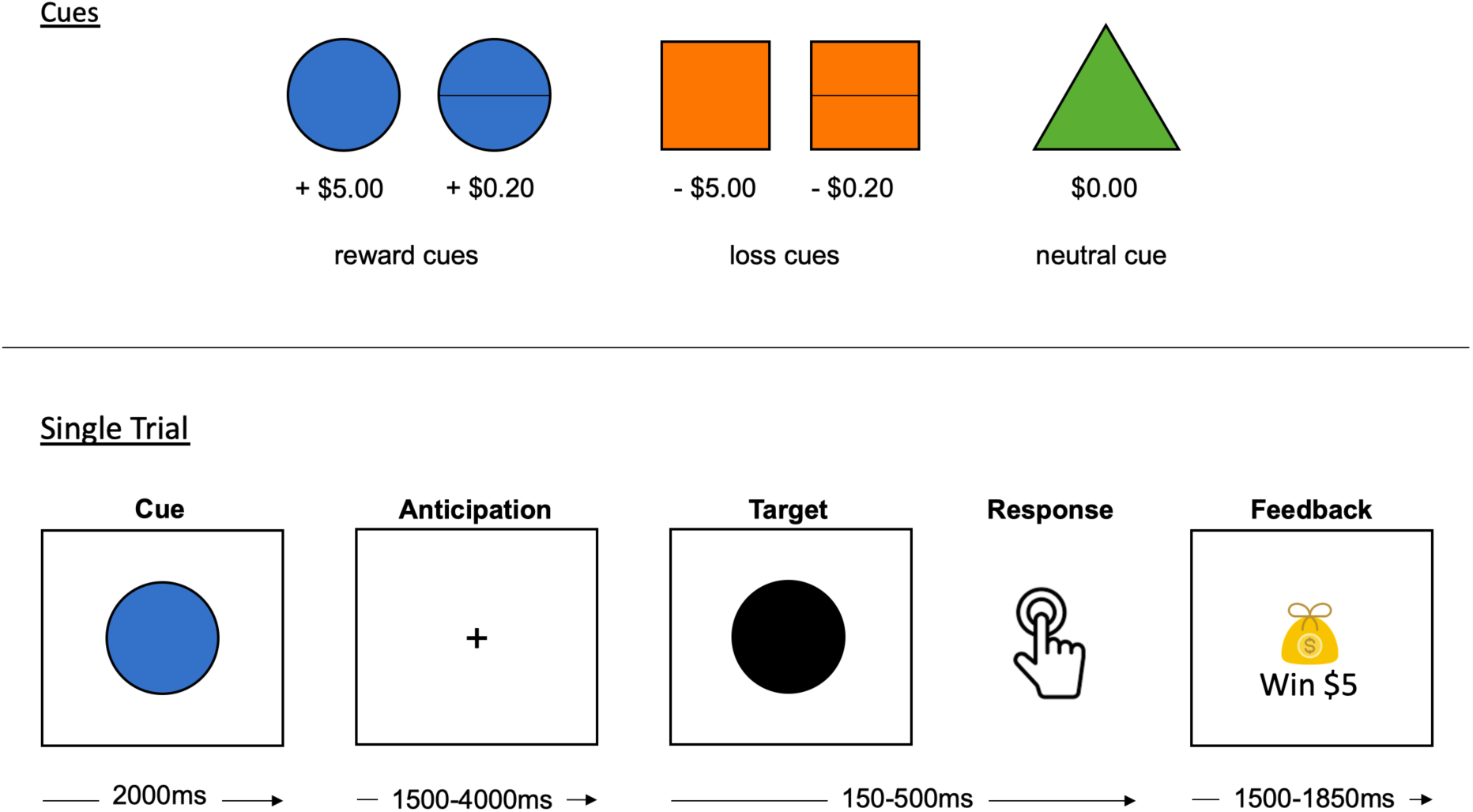
Monetary Incentive Delay Task. Cues and reward/loss associations are presented at the top. A single trial sequence is presented at the bottom.

### Imaging Acquisition and Analysis

Imaging data, detailed in Casey et al. [33], were collected across sites using multichannel coils and multiband echo planar imaging, following a fixed order of scans (localizer, T1- and T2-weighted images, resting state, and diffusion-weighted imaging). Participants completed three tasks (e.g., MID task) in a randomized order. BOLD images were acquired using gradient EPI with standardized parameters.

The ABCD Data Analysis and Informatics Center processed the imaging data, as detailed in Hagler et al. [34]. Regions of interest (ROI) for reward and loss anticipation during the MID task for subcortical regions (bilateral caudate, nucleus accumbens, putamen and thalamus) were derived from the ASEG atlas [35] and cortical regions (dlPFC and vmPFC) were derived from the aparc2009 FreeSurfer atlas [36]. Based on previous literature, the dlPFC beta estimates were derived by combining beta estimates from the middle frontal gyrus and the inferior frontal sulcus [37], and the vmPFC estimates were derived by combining beta estimates from the fronto-marginal gyrus and sulcus, transverse frontopolar gyri and sulci and lateral orbital sulcus [38]. For reasons of statistical power, this study focused on contrasts for “all rewards vs. no money,” “all losses vs. no money” for reward and loss anticipation respectively. Analyses for small and large rewards are presented in the supplements. Average beta coefficients were calculated across all runs.

### Statistical Analysis

We investigated potential group differences in reaction times in respond to rewards and losses separately. For this, we performed robust mixed ANOVAs and robust post hoc tests for significant effects based on trimmed means using a bootstrap method, with average reaction time as the dependent variable, group as the between-subject variable (controls, risk) and time point as the within-subject variable (baseline, 2^nd^ year follow-up).

Similarly, neural differences for anticipation of rewards and losses across the groups were investigated using robust mixed ANOVAs based on trimmed means with bootstrapping, using the beta estimates for the neural activation as the dependent variable, group (controls, risk) as the between-subject variable and region (all ROIs) as the within-subject variable. Mann-Whitney U-tests were applied as post-hoc tests.

Based on the imaging analysis, ROI activations that yielded significant differences between controls and at-risk individuals during anticipation of rewards and losses were included in regression models to predict PPS scores in the 2^nd^ and 3^rd^ year follow-up across all subjects. All models were corrected for age of the PPS score, race, household income and parent’s education.

### General Procedure

Before assessing for group-comparisons, data was inspected for normality of distribution using Shapiro-Wilks test. For non-normal data (Shapiro-Wilks p>0.05), Mann-Whitney U-tests were conducted. RTs and beta weights were inspected for outliers. Values exceeding 1.5 times the interquartile range of the corresponding scores were imputed by the means of the corresponding scores. Before conducting the regression analysis, missing values for household income were imputed using the means of the respective group specific scores.

All statistical analyses were conducted using the R Statistical Software (version 4.1.1) [39]. Data was visualized using the ggplot2 package (version 3.4.2) [40], the ggpubr package (version 0.6.0) [41], and the Hmisc package (version 4.5-0) [42]. Shapiro-Wilks tests, chi-square tests and Mann-Whitney U-tests were conducted with the rstatix package (version 0.7.2) [43]. Robust ANOVAs were conducted using the WRS2 package (version 1.1-4) [44]. Linear regressions were conducted using the stats package (version 4.3.1) [39]. Imputations were performed using the zoo package (version 1.8-12) [45].

## Results

### Behavioural results

Investigating behavioural differences, the robust mixed ANOVA for both anticipation of reward and loss revealed a significant main effect for time point (reward anticipation: F(1,56.62)=84.13, p<0.001; loss anticipation: F(1,57.89)=127.31, p<0.001), but no main effect for group or an interaction effect. Post hoc tests revealed that, across all subjects, average reaction times for anticipation of reward (baseline vs 2^nd^ year follow-up: psihat = 49.26, p<0.001, 95% CI [38.56, 59.96]) as well as for anticipation of loss (baseline vs 2^nd^ year follow-up: psihat = 41.3, p<0.001, 95% CI [33.91, 48.69]) were higher during baseline than 2^nd^ year follow up.

### Imaging results

Investigating differences in reward anticipation related neural activation, robust mixed ANOVA revealed a significant main effect of group (F(1,54.55)=7.13, p=0.01) and region (F(23,38.97)=4.62, p<0.001), but no interaction effect. Robust post hoc tests for group revealed that controls had higher activations than at-risk individuals (controls vs risk: psihat = 0.06, p<0.001, 95% CI [0.05, 0.08]). Hypothesis-based we conducted post hoc tests demonstrating group differences in ROI activations between controls and risk individuals using Mann-Whitney U-tests. Results are displayed in Figure 3. The analysis revealed significant differences in the right caudate at baseline, and in the left and right caudate, the left nucleus accumbens, the right putamen, the right dlPFC, and the left and right vmPFC at the second follow-up.

**Figure 3.**
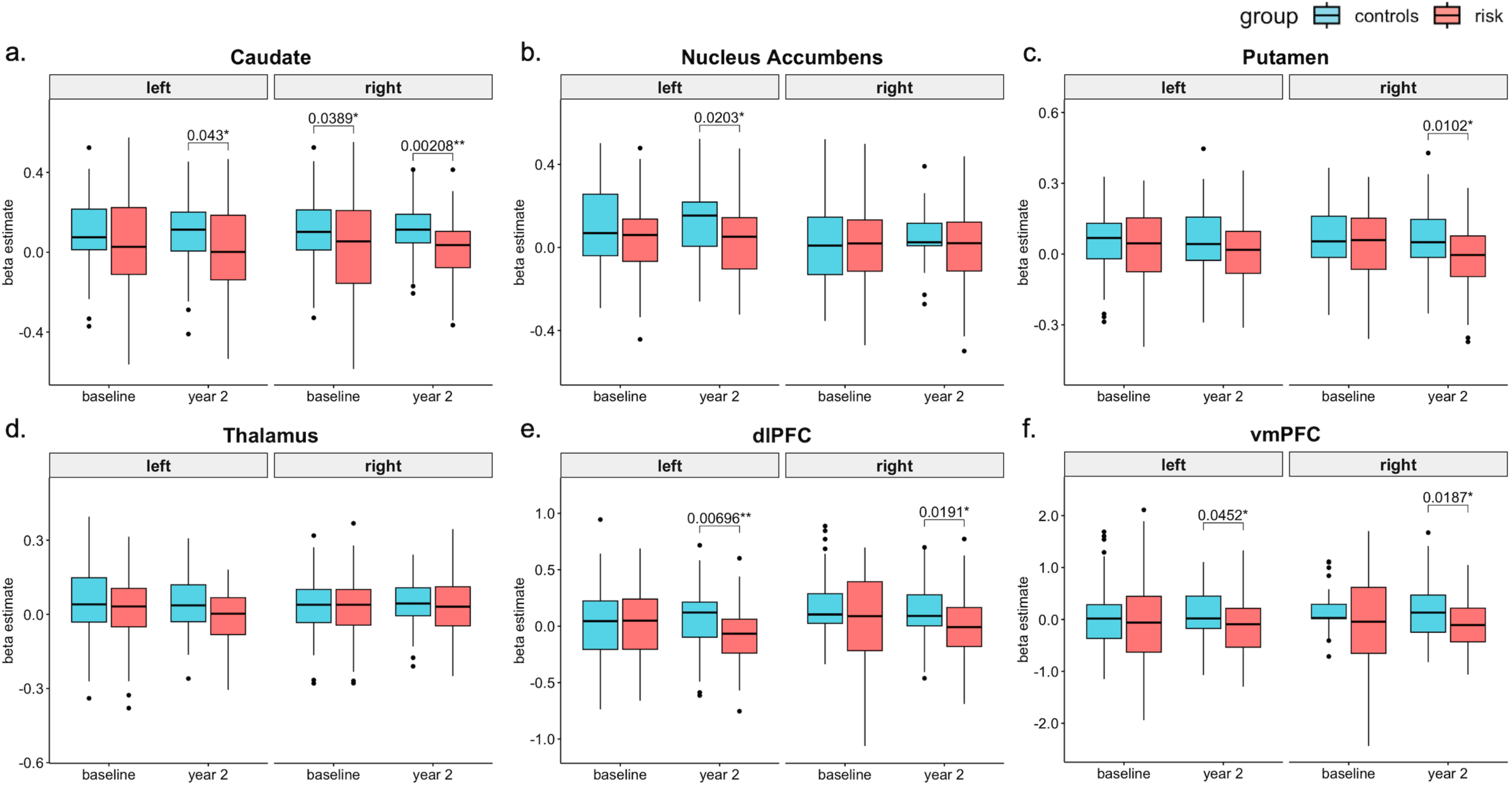
Significant group differences calculated using Mann-Whitney U-tests between control and risk psychosis individuals in brain activations during anticipation of reward in the MID task at baseline and 2^nd^ year follow-up. Note: *: significant differences at p<0.05; **: significant differences at p<0.01. The p-values are not corrected for multiple comparisons due to strictly hypothesis driven analyses.

Robust mixed ANOVA for anticipation of loss revealed a significant main effect of region (F(23,37.93)=1.99, p=0.03), but no group or interaction effect.

### Linear regression results

We explored whether later PPS scores could be predicted based on neural activations which yielded differences across the groups and timepoints. We ran four models:

1. baseline neural activity predicts year 2 PPS scores; 2) baseline and year 2 neural activity predicts year 2 PPS scores; 3) baseline neural activity and year 2 neural activity predicts year 3 PPS scores; and 4) most predictive regions from previous 3 models predict year 3 PPS scores. The equations and the results are presented in Table 2.

**Table 2.**
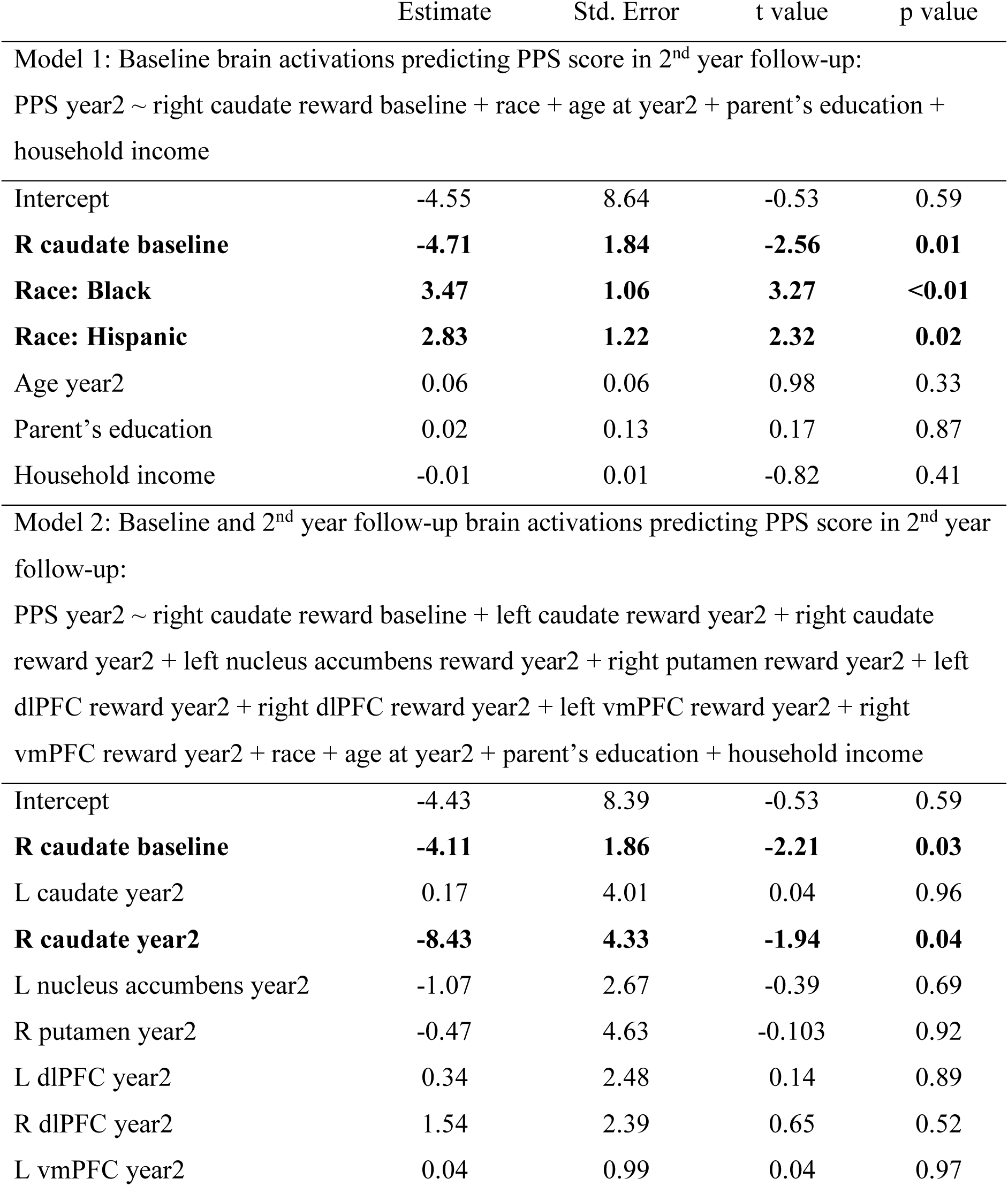
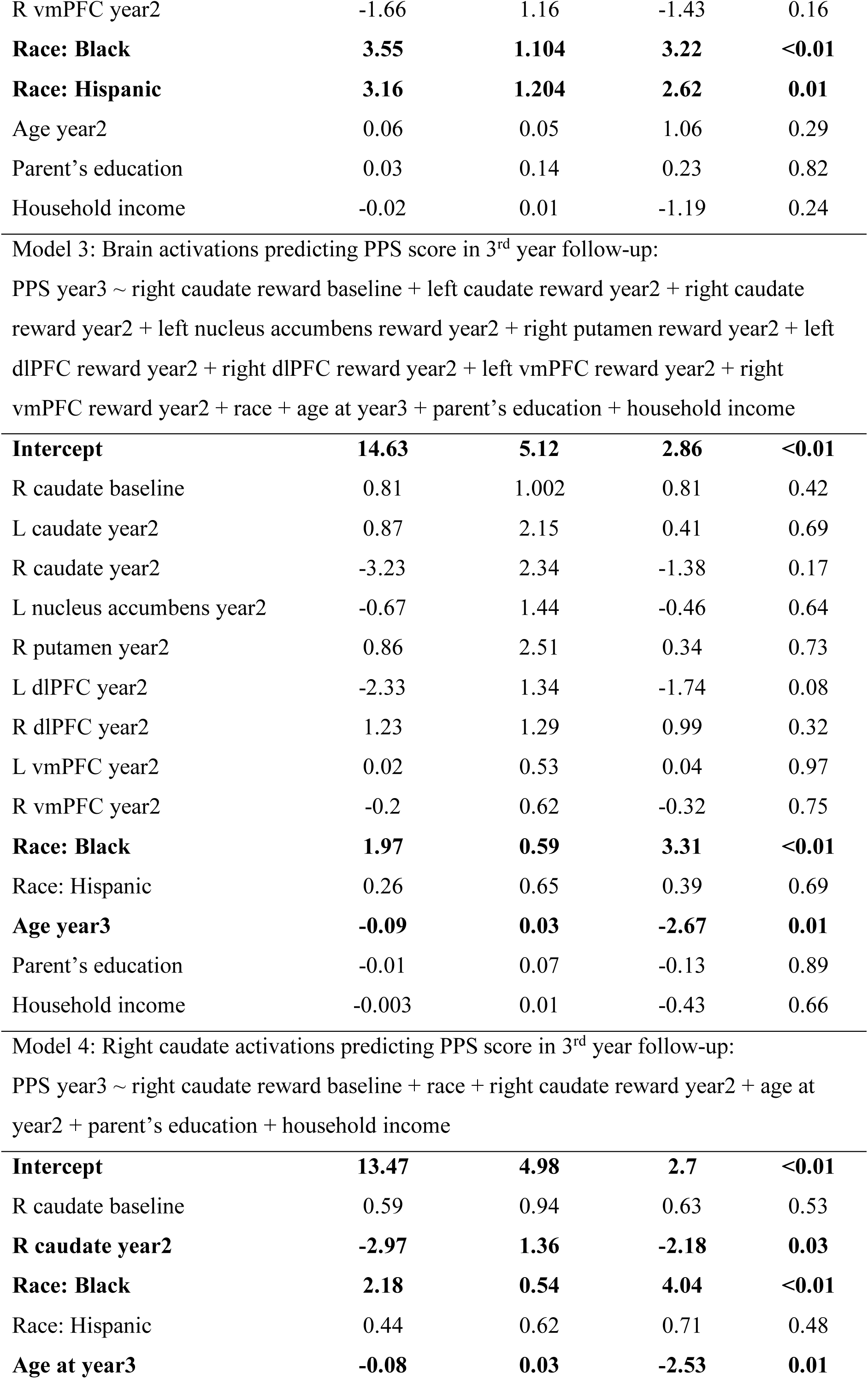
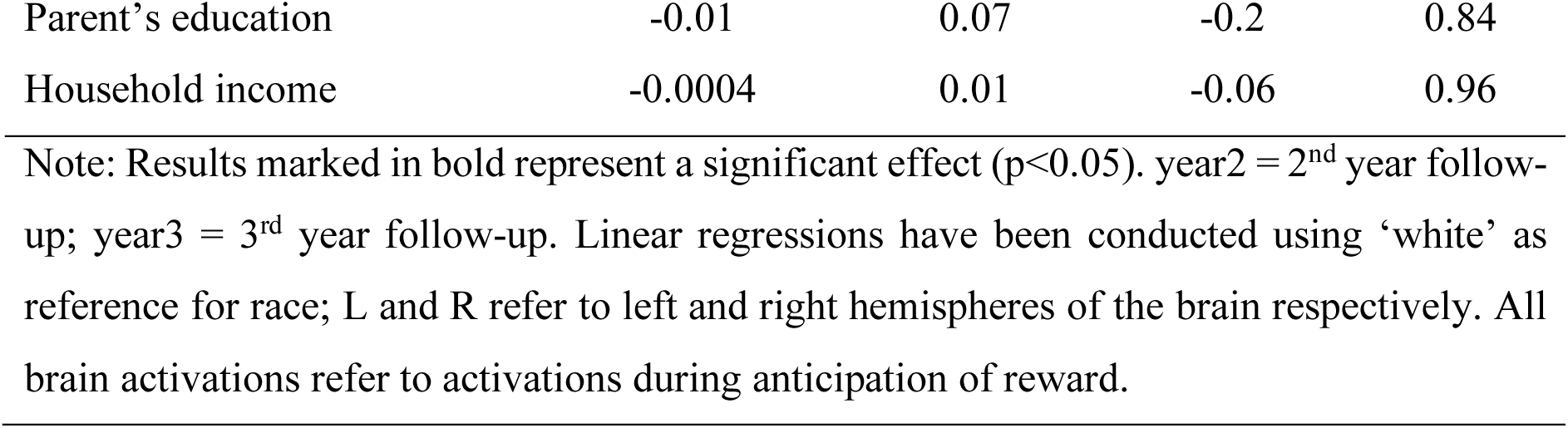
Linear regressions predicting PPS scores in 2^nd^ and 3^rd^ year follow-up.

The linear regression models indicate that right caudate activity at baseline was predictive of PPS score at the 2^nd^ year follow-up, and caudate activity at the 2^nd^ year follow-up was predictive of the PPS score at 2^nd^ year follow-up and also 3^rd^ year follow-up. Interestingly, the covariates revealed that ethnicity was a significant predictor across all models.

## Discussion

In this study, we explored brain alterations by comparing neural correlates of reward and loss anticipation at two timepoints across strictly chosen healthy adolescents and adolescents at-risk for developing groups. Furthermore, we examined whether neural alteration would be predictive of PPS scores at later timepoints. Importantly, we found that brain activations in the striatal and prefrontal cortical regions differed during the anticipation of rewards, consistently showing a hypoactivation. Right caudate activation was significantly lower in at-risk adolescents at baseline. Additionally, activations in the bilateral caudate, right nucleus accumbens, right putamen, bilateral vmPFC, and bilateral dlPFC were significantly lower in at-risk adolescents compared to controls in the 2^nd^ year follow-up. Inmortantly, right caudate activation during reward anticipation at baseline was predictive of PPS scores at the 2^nd^ year follow and right caudate activation during reward anticipation at 2^nd^ year follow-up was predictive of PPS scores at the 2^nd^ year follow-up and 3^rd^ year follow-up, with lower activity linked to higher PPS scores. These findings suggest that right caudate activation potentially plays a crucial role in early psychosis symptoms in adolescents at-risk of psychosis and might be a potential biomarker for early prediction of psychosis.

Our findings of dorsal striatal hypoactivation (right caudate and putamen) in the at-risk group compared to healthy controls align with past findings of reward anticipation to be associated with dorsal striatal regions in older at-risk individuals and first episode psychosis patients [24,25,46]. A recent study by Demro and colleagues [46] investigated reward anticipation in individuals with psychosis and their relatives. In accordance with our findings, their results also revealed a hypoactivation in caudate and putamen, as well as in the nucleus accumbens and the insula in individuals with psychosis compared to controls. Interestingly, the unaffected relatives, however, only showed an intermediate hypoactivation in the caudate [46], indicating the relevance of the dorsal striatum in the pathophysiology of psychosis and its potential role as an intermediate phenotype. Our results extend the existing literature showing that the degree of hypoactivation in the caudate predicted PPS at later timepoints, confirming its relevance in detecting alterations in individuals at increased risk in stages as early as adolescents. Furthermore, the findings indicate the potential predictive power of alterations in the dorsal striatum for risk of psychosis. It is therefore crucial that future studies follow up these alterations in the next ABCD-releases and investigate whether reward anticipation related caudate alterations persist in those at-risk and are predictive of PPS scores (5.0 release contained only about one third of the original sample size and made it impossible to select a risk group across all timepoints). As a study by Sarpal and colleagues [47] demonstrated that antipsychotic treatment with aripiprazole or risperidone not only reduced psychotic symptoms but also altered dorsal striatal connectivity to prefrontal areas, including the OFC, ACC, and DLPFC, persistent caudate hypoactivation during development could serve as an optimal target for early interventions.

We furthermore expected activation differences between the two groups in the nucleus accumbens, supporting our hypothesis, we found hypoactivation in the left nucleus accumbens in at-risk individuals compared to controls. The nucleus accumbens activation, however, was not predictive of PPS scores. The hypoactivation detected in the ventral striatum is in line with past findings. A meta-analysis from multiple MID studies revealed that nucleus accumbens deactivations during reward anticipation were reliably associated with psychosis [23]. Using the ABCD dataset including 6718 adolescents from the baseline time-point, Harju-Seppänen and colleagues [29] also did not observe PPS scores to be associated with nucleus accumbens activations in the MID task during reward anticipation. We assume that the overall endorsement of PPS scores at the baseline timepoint is too low across the entire sample and that only carefully selected risk groups reveal these very early differences. Moreover, a meta-analysis from positron emission tomography studies in schizophrenia [48] observed that dopamine dysregulation, which is responsible for aberrant reward processing in schizophrenia [11,49,50], is greater in the dorsal than in the ventral striatum [48], further supporting our observation of only the activation of the right caudate but not of the nucleus accumbens being predictive of PPS.

In addition to the hypoactivation in the dorsal and ventral striatum, we also found hypoactivation to reward anticipation in the vmPFC and the dlPFC. Based on our and other studies, Figure 4 summarises the areas relevant for alterations in reward anticipation in earliest stages of psychosis. Within this cortico-subcortical reward processing circuit, the thalamus plays a key role by integrating cortical and hypothalamic signals to process cue-reward associations [51], and process salient information to guide attention and behaviour [52]. Thalamic activation during reward anticipation, functions as an ‘alerting signal,’ which then integrates interoceptive information from the insula and relays it to the striatum, where appropriate action responses are selected [52]. In both at-risk adults and adolescents, smaller thalamic nuclei have been observed [53,54]. Additionally, structural and functional thalamocortical dysconnectivity is a consistent finding in genetic and clinical risk individuals [55]. However, contrary to previous research, we did not observe significant differences in thalamic activation between our study groups, which may be attributed to the younger age of the participants. Future studies, particularly those utilizing the 4th-year follow-up of the ABCD data, should further investigate whether such activation differences emerge as the participants grow older.

**Figure 4.**
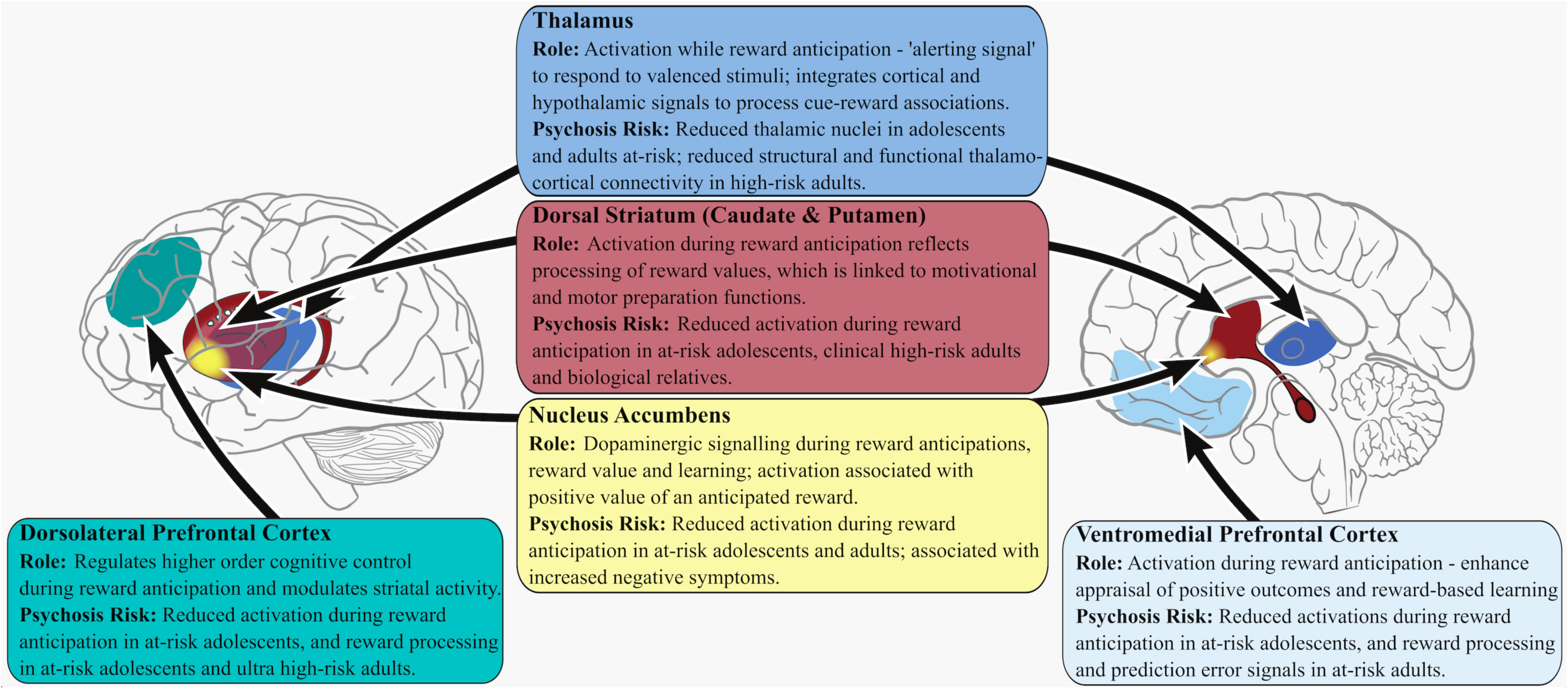
Role of regions during reward anticipation and observed differences in at-risk psychosis individuals.

Likewise, neuroimaging studies have consistently demonstrated robust activation of the dorsal striatum during reward anticipation [65,66]. This activation signals the expected value of rewards and prepares appropriate motivational and motor responses [66,67]. Individuals at a clinical high risk for psychosis show diminished activations in the putamen during reward anticipation [68]. Additionally, biological relatives of adults with a history of psychosis exhibit reduced caudate activation during reward anticipation compared to healthy controls in [46]. Furthermore, adolescents with PLEs show reduced activation in the dorsal striatum during reward processing [69]. Along with the dorsal striatum, the ventral striatum—particularly the nucleus accumbens—plays a crucial role in enhancing motivation and reinforcing reward-associated behaviours through dopaminergic signalling, playing a crucial role in reward anticipation and experience [70]. The nucleus accumbens is also involved in processing the subjective value of future rewards, aiding in reward learning [65,66,71], and is specifically associated with the positive value of anticipated rewards [72]. However, at-risk individuals show reduced activations in the nucleus accumbens [23,73], which has been linked to increased negative symptoms in these individuals [11]. Striatal deactivations during reward anticipation observed in our study may therefore reflect early signs of altered motivational and reward mechanisms, and might contribute to the development of negative symptoms or decreased goal-directed behaviours with age.

In contrast to the thalamus and striatum, the dlPFC plays a critical role regulating higher order cognitive control [56,57], along with modulating striatal activity during reward anticipation [58,59]. Adolescents with PLEs [60] and adult at-risk individuals [61] demonstrate reduced dLPFC activation during reward processing compared to healthy controls. On the other hand, the vmPFC is primarily involved in regulating the subjective value of the reward [25,56,62], by modulating physiological responses such as heart rate variability [62]. Recent findings suggest that vmPFC activation enhances outcome anticipation and reward-based learning by inducing a positive bias in the reward system [63]. However, in at-risk individuals, the vmPFC shows reduced activations, particularly in response to rewards and reward-prediction error signals [64]. Reduced activation in the dlPFC and vmPFC during reward anticipation in our study might reflect early impairments in cognitive control and subjective reward valuation, which could potentially contribute to difficulties in reward-based learning in the future.

Although we found differences across multiple subcortical and cortical reward processing areas, only those in the associated dorsal striatum are predictive of PPS scores at later time-points. Thus, our findings are in line with the hypothesis that subcortical alterations of reward processing are indicators of the manifestation of the illness, and they precede cortical alterations [11].

While we report differences in neural responses to reward anticipation, the behavioural performance in the risk group was similar to healthy individuals, supporting previous research [26,74]. This is in line with other studies showing that neurobiological abnormalities are present before psychosis onset [75,76]. Other studies, however, show that computational parameters of reward processing may be used to unravel early dysfunctions [16,17,77] and may enhance psychosis prediction [78]. Therefore, future studies should apply computational modelling to reward anticipation tasks such as the MID task to explore whether these mathematical approaches reveal latent constructs that show neuropsychological differences at these early stages, and that might further improve the predictive potential of these dysfunctions.

Furthermore, regression analysis suggested that higher PPS scores were associated with being Black and/or Hispanics but not White. This is consistent with previous literature indicating that individuals belonging to these ethnicities were frequently diagnosed with schizophrenia and reported more psychotic experiences compared to other racial groups [79–81]. This finding holds significant implications for policymakers and public health authorities, as it underscores the urgent need for systemic changes and enhanced social support for these communities. Addressing these disparities may not only improve diagnostic accuracy and access to care, but also mitigate the disproportionate burden of psychotic disorders experienced by Black and Hispanic individuals. Such changes require coordinated efforts to reform healthcare delivery and expand social services, rather than relying solely on medical interventions.

Moreover, our regression analysis also observed a decline in PPS scores with age in the 3^rd^ time-point, which is also consistent with past meta-analysis showing psychotic symptoms to be more prevalent in younger compared to older adolescents [82]. They observed that individuals with prodromal risk syndromes had a higher prevalence of co-occurring non-psychotic psychiatric disorders [83]. They suggested that research conducted during late adolescence, when the risk of psychosis peaks, might not reveal a similarly high prevalence of prodromal risk syndromes as observed in the younger population. Given our findings, it will be useful to monitor PPS scores and the rate of conversion to psychosis using a longitudinal design in the future.

Taken together, right caudate activation seems to be a primary predictive neural marker of psychosis risk. Future studies should aim to investigate the associations between PPS scores and caudate activations in a longitudinal setting using a larger sample size of adolescents at a high risk for psychosis, potentially also with familial history of schizophrenia. It will also be interesting to monitor if these associations are able to predict the onset of psychosis over time.

### Conclusion

This study affirms that alterations in brain activations already manifest in early adolescents at an elevated risk of psychosis, even as their behavioural responses remain unaffected. Importantly, lower activations in the right caudate during reward anticipation, were found to be predictive of high psychosis symptoms at a later time-point. These findings hint at a potential role of right caudate deactivations as a predictive biomarker for psychosis. Future studies should seek to validate these findings in ultra-high-risk children with a family history of psychotic disorders.

## Data Availability

Data used in the preparation of this article were obtained from the Adolescent Brain Cognitive DevelopmentSM (ABCD) Study (https://abcdstudy.org), held in the NIMH Data Archive (NDA).

## Acknowledgements

Data used in the preparation of this article were obtained from the Adolescent Brain Cognitive Development^SM^ (ABCD) Study (https://abcdstudy.org), held in the NIMH Data Archive (NDA). This is a multisite, longitudinal study designed to recruit more than 10 000 children age 9–10 and follow them over 10 years into early adulthood. The ABCD Study is supported by the National Institutes of Health and additional federal partners under award numbers

U01DA041022, U01DA041028, U01DA041048, U01DA041089, U01DA041106, U01DA041117, U01DA041120, U01DA041134, U01DA041148, U01DA041156, U01DA041174, U24DA041123, U24DA041147, U01DA041093, and U01DA041025. A full list of supporters is available at https://abcdstudy.org/federal-partners.html. A listing of participating sites and a complete listing of the study investigators can be found at https://abcdstudy.org/Consortium_Members.pdf. ABCD consortium investigators designed and implemented the study and/or provided data but did not necessarily participate in analysis or writing of this report. This manuscript reflects the views of the authors and may not reflect the opinions or views of the NIH or ABCD consortium investigators. The ABCD data repository grows and changes over time. The ABCD data used in this report came from doi:10.15154/1523041.

Furthermore, we would like to thank Viola von Heyden for her contribution to this project.

## Conflicts of Interest

None of the authors report any conflicts of interest.

## Notes

### Competing Interest Statement

The authors have declared no competing interest.

### Funding Statement

This study did not receive any funding.

### Author Declarations

We used data from the Adolescent Brain and Cognitive Development study.

## References

[1] Saxena A, Dodell-Feder D. Environmental Risk Factors for Childhood Psychotic-Like Experiences in the Adolescent Brain Cognitive Development (ABCD) Datase 2020. 10.31234/osf.io/z546j.

[2] Legge SE, Santoro ML, Periyasamy S, Okewole A, Arsalan A, Kowalec K. Genetic architecture of schizophrenia: a review of major advancements. Psychological Medicine 2021;51:2168–77. 10.1017/S0033291720005334.

[3] Tanskanen A, Tiihonen J, Taipale H. Mortality in schizophrenia: 30-year nationwide follow-up study. Acta Psychiatrica Scandinavica 2018;138:492–9. 10.1111/acps.12913.

[4] Correll CU, Galling B, Pawar A, Krivko A, Bonetto C, Ruggeri M, et al. Comparison of Early Intervention Services vs Treatment as Usual for Early-Phase Psychosis: A Systematic Review, Meta-analysis, and Meta-regression. JAMA Psychiatry 2018;75:555–65. 10.1001/jamapsychiatry.2018.0623.

[5] Schmidt SJ, Schultze-Lutter F, Schimmelmann BG, Maric NP, Salokangas RKR, Riecher-Rössler A, et al. EPA guidance on the early intervention in clinical high risk states of psychoses. European Psychiatry 2015;30:388–404. 10.1016/j.eurpsy.2015.01.013.

[6] Guo JY, Niendam TA, Auther AM, Carrión RE, Cornblatt BA, Ragland JD, et al. Predicting psychosis risk using a specific measure of cognitive control: a 12-month longitudinal study. Psychological Medicine 2020;50:2230–9. DOI: 10.1017/S0033291719002332.

[7] Rosen M, Betz LT, Schultze-Lutter F, Chisholm K, Haidl TK, Kambeitz-Ilankovic L, et al. Towards clinical application of prediction models for transition to psychosis: a systematic review and external validation study in the PRONIA sample. Neuroscience & Biobehavioral Reviews 2021;125:478–92.

[8] Sullivan SA, Kounali D, Cannon M, David AS, Fletcher PC, Holmans P, et al. A population-based cohort study examining the incidence and impact of psychotic experiences from childhood to adulthood, and prediction of psychotic disorder. American Journal of Psychiatry 2020;177:308–17.

[9] Hochberger W, Thomas M, Joshi Y, Swerdlow N, Braff D, Gur R, et al. Deviation from expected cognitive ability is a core cognitive feature of schizophrenia related to neurophysiologic, clinical and psychosocial functioning. Schizophrenia Research 2020;215:300–7.

[10] Moran EK, Culbreth AJ, Kandala S, Barch DM. From neuroimaging to daily functioning: A multimethod analysis of reward anticipation in people with schizophrenia. Journal of Abnormal Psychology 2019;128:723.

[11] Kesby JP, Murray GK, Knolle F. Neural Circuitry of Salience and Reward Processing in Psychosis. Biological Psychiatry Global Open Science 2023;3:33–46. 10.1016/j.bpsgos.2021.12.003.

[12] Chung YS, Barch DM. Frontal-Striatum Dysfunction During Reward Processing: Relationships to Amotivation in Schizophrenia. J Abnorm Psychol 2016;125:453–69. 10.1037/abn0000137.

[13] Heinz A. Dopaminergic dysfunction in alcoholism and schizophrenia - Psychopathological and behavioral correlates. European Psychiatry 2002. 10.1016/S0924-9338(02)00628-4.

[14] Kapur S. Psychosis as a state of aberrant salience: A framework linking biology, phenomenology, and pharmacology in schizophrenia. American Journal of Psychiatry 2003;160:13–23. 10.1176/appi.ajp.160.1.13.

[15] Baker A, Suetani S, Cosgrove P, Siskind D, Murray GK, Scott JG, et al. Reversal learning in those with early psychosis features contingency-dependent changes in loss response and learning. Cognitive Neuropsychiatry 2023:1–19.

[16] Knolle F, Sterner EF, Moutoussis M, Adams RA, Griffin JD, Haarsma J, et al. Action selection in early stages of psychosis: an active inference approach. JOURNAL OF PSYCHIATRY & NEUROSCIENCE 2023;ahead.

[17] Montagnese M, Knolle F, Haarsma J, Griffin JD, Richards A, Vertes PE, et al. Reinforcement learning as an intermediate phenotype in psychosis? Deficits sensitive to illness stage but not associated with polygenic risk of schizophrenia in the general population. Schizophrenia Research 2020.

[18] Brandl F, Knolle F, Avram M, Leucht C, Yakushev I, Priller J, et al. Negative symptoms, striatal dopamine and model-free reward decision-making in schizophrenia. Brain 2023;146:767–77.

[19] Ermakova AO, Knolle F, Justicia A, Bullmore ET, Jones PB, Robbins TW, et al. Abnormal reward prediction-error signalling in antipsychotic naive individuals with first-episode psychosis or clinical risk for psychosis. Neuropsychopharmacol 2018;43:1691–9. 10.1038/s41386-018-0056-2.

[20] Haarsma J, Fletcher PC, Griffin JD, Taverne HJ, Ziauddeen H, Spencer TJ, et al. Precision weighting of cortical unsigned prediction error signals benefits learning, is mediated by dopamine, and is impaired in psychosis. Molecular Psychiatry 2020. 10.1038/s41380-020-0803-8.

[21] Murray G, Corlett P, Clark L, Pessiglione M, Blackwell A, Honey G, et al. Substantia nigra / ventral tegmental reward prediction error disruption in psychosis. Molecular Psychiatry 2008;13:1–18. 10.1038/sj.mp.4002058.Substantia.

[22] Knutson B, Heinz A. Probing Psychiatric Symptoms with the Monetary Incentive Delay Task. Biological Psychiatry 2015;77:418–20. 10.1016/j.biopsych.2014.12.022.

[23] Radua J, Schmidt A, Borgwardt S, Heinz A, Schlagenhauf F, McGuire P, et al. Ventral Striatal Activation During Reward Processing in Psychosis: A Neurofunctional Meta-Analysis. JAMA Psychiatry 2015;72:1243–51. 10.1001/jamapsychiatry.2015.2196.

[24] Subramaniam K, Hooker CI, Biagianti B, Fisher M, Nagarajan S, Vinogradov S. Neural signal during immediate reward anticipation in schizophrenia: Relationship to real-world motivation and function. NeuroImage: Clinical 2015;9:153–63. 10.1016/j.nicl.2015.08.001.

[25] Zeng J, Yan J, Cao H, Su Y, Song Y, Luo Y, et al. Neural substrates of reward anticipation and outcome in schizophrenia: a meta-analysis of fMRI findings in the monetary incentive delay task. Transl Psychiatry 2022;12:1–14. 10.1038/s41398-022-02201-8.

[26] Michielse S, Lange I, Bakker J, Goossens L, Verhagen S, Papalini S, et al. Reward anticipation in individuals with subclinical psychotic experiences: A functional MRI approach. European Neuropsychopharmacology 2019;29:1374–85. 10.1016/j.euroneuro.2019.10.002.

[27] Nielsen MØ, Rostrup E, Wulff S, Bak N, Lublin H, Kapur S, et al. Alterations of the Brain Reward System in Antipsychotic Naïve Schizophrenia Patients. Biological Psychiatry 2012;71:898–905. 10.1016/j.biopsych.2012.02.007.

[28] Tangmose K, Rostrup E, Bojesen KB, Sigvard A, Jessen K, Johansen LB, et al. Reward disturbances in antipsychotic-naïve patients with first-episode psychosis and their association to glutamate levels. Psychol Med 2023;53:1629–38. 10.1017/S0033291721003305.

[29] Harju-Seppänen J, Irizar H, Bramon E, Blakemore S-J, Mason L, Bell V. Reward Processing in Children With Psychotic-Like Experiences. Schizophrenia Bulletin Open 2022;3:sgab054. 10.1093/schizbullopen/sgab054.

[30] Knutson B, Westdorp A, Kaiser E, Hommer D. FMRI visualization of brain activity during a monetary incentive delay task. Neuroimage 2000;12:20–7. 10.1006/nimg.2000.0593.

[31] Karcher NR, Barch DM, Avenevoli S, Savill M, Huber RS, Simon TJ, et al. Assessment of the Prodromal Questionnaire–Brief Child Version for Measurement of Self-reported Psychoticlike Experiences in Childhood. JAMA Psychiatry 2018;75:853–61. 10.1001/jamapsychiatry.2018.1334.

[32] Loewy RL, Pearson R, Vinogradov S, Bearden CE, Cannon TD. Psychosis risk screening with the Prodromal Questionnaire — Brief Version (PQ-B). Schizophrenia Research 2011;129:42–6. 10.1016/j.schres.2011.03.029.

[33] Casey BJ, Cannonier T, Conley MI, Cohen AO, Barch DM, Heitzeg MM, et al. The Adolescent Brain Cognitive Development (ABCD) study: Imaging acquisition across 21 sites. Developmental Cognitive Neuroscience 2018;32:43–54. 10.1016/j.dcn.2018.03.001.

[34] Hagler Jr DJ, Hatton S, Cornejo MD, Makowski C, Fair DA, Dick AS, et al. Image processing and analysis methods for the Adolescent Brain Cognitive Development Study. Neuroimage 2019;202:116091.

[35] Fischl B, Salat DH, Busa E, Albert M, Dieterich M, Haselgrove C, et al. Whole brain segmentation: automated labeling of neuroanatomical structures in the human brain. Neuron 2002;33:341–55.

[36] Destrieux C, Fischl B, Dale A, Halgren E. Automatic parcellation of human cortical gyri and sulci using standard anatomical nomenclature. NeuroImage 2010;53:1–15. 10.1016/j.neuroimage.2010.06.010.

[37] Chaarani B, Hahn S, Allgaier N, Adise S, Owens MM, Juliano AC, et al. Baseline brain function in the preadolescents of the ABCD Study. Nat Neurosci 2021;24:1176–86. 10.1038/s41593-021-00867-9.

[38] Fariña A, Rojek-Giffin M, Gross J, De Dreu CKW. Social preferences correlate with cortical thickness of the orbito-frontal cortex. Soc Cogn Affect Neurosci 2021;16:1191–203. 10.1093/scan/nsab074.

[39] R Core Team. R: A Language and Environment for Statistical Computing. Vienna, Austria: R Foundation for Statistical Computing; 2021.

[40] Wickham H. ggplot2: Elegant Graphics for Data Analysis. Springer-Verlag New York; 2016.

[41] Kassambara A. ggpubr: “ggplot2” Based Publication Ready Plots. 2020.

[42] Jr FEH, Dupont with contributions from C, others many. Hmisc: Harrell Miscellaneous. 2021.

[43] Singmann H, Bolker B, Westfall J, Aust F, Ben-Shachar MS. afex: Analysis of Factorial Experiments. 2021.

[44] Mair P, Wilcox R. Robust statistical methods in R using the WRS2 package. Behaviour Research Methods 2019;52:464–88.

[45] Zeileis A, Grothendieck G. zoo: S3 Infrastructure for Regular and Irregular Time Series. Journal of Statistical Software 2005;14:1–27. 10.18637/jss.v014.i06.

[46] Demro C, Lahud E, Burton PC, Purcell JR, Simon JJ, Sponheim SR. Reward anticipation-related neural activation following cued reinforcement in adults with psychotic psychopathology and biological relatives. Psychological Medicine 2024;54:1441–51. 10.1017/S0033291723003343.

[47] Sarpal DK, Robinson DG, Lencz T, Argyelan M, Ikuta T, Karlsgodt K, et al. Antipsychotic Treatment and Functional Connectivity of the Striatum in First-Episode Schizophrenia. JAMA Psychiatry 2015;72:5–13. 10.1001/jamapsychiatry.2014.1734.

[48] McCutcheon R, Beck K, Jauhar S, Howes OD. Defining the Locus of Dopaminergic Dysfunction in Schizophrenia: A Meta-analysis and Test of the Mesolimbic Hypothesis. Schizophr Bull 2018;44:1301–11. 10.1093/schbul/sbx180.

[49] Howes OD, Kapur S. The dopamine hypothesis of schizophrenia: version III--the final common pathway. Schizophr Bull 2009;35:549–62. 10.1093/schbul/sbp006.

[50] Matthews M, Bondi C, Torres G, Moghaddam B. Reduced Presynaptic Dopamine Activity in Adolescent Dorsal Striatum. Neuropsychopharmacol 2013;38:1344–51. 10.1038/npp.2013.32.

[51] Otis JM, Zhu M, Namboodiri VM, Cook CA, Kosyk O, Matan AM, et al. Paraventricular thalamus projection neurons integrate cortical and hypothalamic signals for cue-reward processing. Neuron 2019;103:423–31.

[52] Cho YT, Fromm S, Guyer AE, Detloff A, Pine DS, Fudge JL, et al. Nucleus accumbens, thalamus and insula connectivity during incentive anticipation in typical adults and adolescents,. Neuroimage 2013;0:508–21. 10.1016/j.neuroimage.2012.10.013.

[53] Benoit LJ, Canetta S, Kellendonk C. Thalamocortical Development: A Neurodevelopmental Framework for Schizophrenia. Biol Psychiatry 2022;92:491–500. 10.1016/j.biopsych.2022.03.004.

[54] Huang AS, Rogers BP, Sheffield JM, Jalbrzikowski ME, Anticevic A, Blackford JU, et al. Thalamic nuclei volumes in psychotic disorders and in youths with psychosis spectrum symptoms. American Journal of Psychiatry 2020;177:1159–67.

[55] Steullet P. Thalamus-related anomalies as candidate mechanism-based biomarkers for psychosis. Schizophrenia Research 2020;226:147–57. 10.1016/j.schres.2019.05.027.

[56] Nejati V, Majdi R, Salehinejad MA, Nitsche MA. The role of dorsolateral and ventromedial prefrontal cortex in the processing of emotional dimensions. Sci Rep 2021;11:1971. 10.1038/s41598-021-81454-7.

[57] Wilson RP, Colizzi M, Bossong MG, Allen P, Kempton M, Abe N, et al. The Neural Substrate of Reward Anticipation in Health: A Meta-Analysis of fMRI Findings in the Monetary Incentive Delay Task. Neuropsychol Rev 2018;28:496–506. 10.1007/s11065-018-9385-5.

[58] Becker A, Kirsch M, Gerchen MF, Kiefer F, Kirsch P. Striatal activation and frontostriatal connectivity during non-drug reward anticipation in alcohol dependence. Addiction Biology 2017;22:833–43. 10.1111/adb.12352.

[59] Staudinger MR, Erk S, Walter H. Dorsolateral Prefrontal Cortex Modulates Striatal Reward Encoding during Reappraisal of Reward Anticipation. Cerebral Cortex 2011;21:2578–88. 10.1093/cercor/bhr041.

[60] Bourque J, Spechler PA, Potvin S, Whelan R, Banaschewski T, Bokde AL, et al. Functional neuroimaging predictors of self-reported psychotic symptoms in adolescents. American Journal of Psychiatry 2017;174:566–75.

[61] Fusar-Poli P, Perez J, Broome M, Borgwardt S, Placentino A, Caverzasi E, et al. Neurofunctional correlates of vulnerability to psychosis: A systematic review and meta-analysis. Neuroscience & Biobehavioral Reviews 2007;31:465–84. 10.1016/j.neubiorev.2006.11.006.

[62] Motzkin JC, Philippi CL, Wolf RC, Baskaya MK, Koenigs M. Ventromedial Prefrontal Cortex Lesions Alter Neural and Physiological Correlates of Anticipation. J Neurosci 2014;34:10430–7. 10.1523/JNEUROSCI.1446-14.2014.

[63] Rehbein MA, Kroker T, Winker C, Ziehfreund L, Reschke A, Bölte J, et al. Non-invasive stimulation reveals ventromedial prefrontal cortex function in reward prediction and reward processing. Frontiers in Neuroscience 2023;17:1219029.

[64] Millman ZB, Gallagher K, Demro C, Schiffman J, Reeves GM, Gold JM, et al. Evidence of reward system dysfunction in youth at clinical high-risk for psychosis from two event-related fMRI paradigms. Schizophrenia Research 2020;226:111–9. 10.1016/j.schres.2019.03.017.

[65] Carruzzo F, Giarratana AO, del Puppo L, Kaiser S, Tobler PN, Kaliuzhna M. Neural bases of reward anticipation in healthy individuals with low, mid, and high levels of schizotypy. Sci Rep 2023;13:9953. 10.1038/s41598-023-37103-2.

[66] Oldham S, Murawski C, Fornito A, Youssef G, Yücel M, Lorenzetti V. The anticipation and outcome phases of reward and loss processing: A neuroimaging meta-analysis of the monetary incentive delay task. Human Brain Mapping 2018;39:3398–418. 10.1002/hbm.24184.

[67] Balleine BW, Delgado MR, Hikosaka O. The Role of the Dorsal Striatum in Reward and Decision-Making. J Neurosci 2007;27:8161–5. 10.1523/JNEUROSCI.1554-07.2007.

[68] Zeng J, Yan J, You L, Liao T, Luo Y, Cheng B, et al. A Meta-Analysis of Neural Correlates of Reward Anticipation in Individuals at Clinical Risk for Schizophrenia. International Journal of Neuropsychopharmacology 2023;26:280–93. 10.1093/ijnp/pyad009.

[69] Papanastasiou E, Mouchlianitis E, Joyce DW, McGuire P, Banaschewski T, Bokde ALW, et al. Examination of the Neural Basis of Psychoticlike Experiences in Adolescence During Reward Processing. JAMA Psychiatry 2018;75:1043–51. 10.1001/jamapsychiatry.2018.1973.

[70] Klawonn AM, Malenka RC. Nucleus Accumbens Modulation in Reward and Aversion. Cold Spring Harbor Symposia on Quantitative Biology 2018;83:119. 10.1101/sqb.2018.83.037457.

[71] Galvan A, Hare TA, Davidson M, Spicer J, Glover G, Casey BJ. The Role of Ventral Frontostriatal Circuitry in Reward-Based Learning in Humans. J Neurosci 2005;25:8650–6. 10.1523/JNEUROSCI.2431-05.2005.

[72] Knutson B, Adams CM, Fong GW, Hommer D. Anticipation of Increasing Monetary Reward Selectively Recruits Nucleus Accumbens. J Neurosci 2001;21:RC159–RC159. 10.1523/JNEUROSCI.21-16-j0002.2001.

[73] Karcher NR, Hua JPY, Kerns JG. Striatum-related functional activation during reward-versus punishment-based learning in psychosis risk. Neuropsychopharmacology 2019;44:1967–74. 10.1038/s41386-019-0455-z.

[74] Wotruba D, Heekeren K, Michels L, Buechler R, Simon JJ, Theodoridou A, et al. Symptom dimensions are associated with reward processing in unmedicated persons at risk for psychosis. Front Behav Neurosci 2014;8. 10.3389/fnbeh.2014.00382.

[75] Borgwardt SJ, Riecher-Rössler A, Dazzan P, Chitnis X, Aston J, Drewe M, et al. Regional Gray Matter Volume Abnormalities in the At Risk Mental State. Biological Psychiatry 2007;61:1148–56. 10.1016/j.biopsych.2006.08.009.

[76] Fusar-Poli P, Howes OD, Allen P, Broome M, Valli I, Asselin M-C, et al. Abnormal prefrontal activation directly related to pre-synaptic striatal dopamine dysfunction in people at clinical high risk for psychosis. Mol Psychiatry 2011;16:67–75. 10.1038/mp.2009.108.

[77] Haarsma J, Knolle F, Griffin JD, Taverne H, Mada M, Goodyer IM, et al. Influence of prior beliefs on perception in early psychosis: Effects of illness stage and hierarchical level of belief. Journal of Abnormal Psychology 2020;129:581.

[78] Gold JM, Corlett PR, Strauss GP, Schiffman J, Ellman LM, Walker EF, et al. Enhancing psychosis risk prediction through computational cognitive neuroscience. Schizophrenia Bulletin 2020;46:1346–52.

[79] Cohen CI, Marino L. Racial and Ethnic Differences in the Prevalence of Psychotic Symptoms in the General Population. PS 2013;64:1103–9. 10.1176/appi.ps.201200348.

[80] Posternak MA, Zimmerman M. Elevated Rates of Psychosis Among Treatment-Seeking Hispanic Patients With Major Depression. The Journal of Nervous and Mental Disease 2005;193:66. 10.1097/01.nmd.0000149222.02563.be.

[81] Schwartz RC, Blankenship DM. Racial disparities in psychotic disorder diagnosis: A review of empirical literature. World J Psychiatry 2014;4:133–40. 10.5498/wjp.v4.i4.133.

[82] Kelleher I, Connor D, Clarke MC, Devlin N, Harley M, Cannon M. Prevalence of psychotic symptoms in childhood and adolescence: a systematic review and meta-analysis of population-based studies. Psychol Med 2012;42:1857–63. 10.1017/S0033291711002960.

[83] Kelleher I, Murtagh A, Molloy C, Roddy S, Clarke MC, Harley M, et al. Identification and Characterization of Prodromal Risk Syndromes in Young Adolescents in the Community: A Population-Based Clinical Interview Study. Schizophr Bull 2012;38:239–46. 10.1093/schbul/sbr164.

