## Supplementary Material for "Right caudate deactivation during reward anticipation predicts elevated risk for psychosis in adolescents"

*corresponding author:

Dr. Franziska Knolle

**Differences in neural activation during reward anticipation: Large reward vs no money baseline**

Investigating differences in neural activation during reward anticipation at baseline for the contrast “large reward vs. no money”, robust mixed ANOVA revealed a significant main effect of region (F(11,38.26)=2.35, p=0.02), but no main effect for group or interaction effect.

Group differences in ROI activations at baseline between controls and risk individuals using Mann-Whitney U-tests for the contrast “large reward vs. no money” are displayed in Supplementary Table 1.

**Supplementary Table 1.** Group differences at baseline (large reward vs. no money)

| ROI | Group 1 | Group 2 | W statistic | p-value |
| --- | --- | --- | --- | --- |
| Left Caudate | controls | risk | 1306 | 0.12 |
| **Right Caudate** | **controls** | **risk** | **1387** | **0.03** |
| **Left Nucleus Accumbens** | **controls** | **risk** | **1398** | **0.03** |
| Right Nucleus Accumbens | controls | risk | 1031 | 0.84 |
| Left Putamen | controls | risk | 1248 | 0.24 |
| Right Putamen | controls | risk | 1102 | 0.27 |
| Left Thalamus | controls | risk | 1231 | 0.18 |
| Right Thalamus | controls | risk | 1197 | 0.28 |
| Left dlPFC | controls | risk | 1254 | 0.18 |
| Right dlPFC | controls | risk | 1142 | 0.27 |
| Left vmPFC | controls | risk | 1299 | 0.14 |
| Right vmPFC | controls | risk | 1091 | 0.51 |
| Note: Group differences are calculated using Mann-Whitney U-tests. Results marked in bold represent a significant effect (p<0.05). | | | | |

**Differences in neural activation during reward anticipation: Large reward vs no money year2**

Investigating differences in neural activation during reward anticipation at 2^nd^ year follow-up for the contrast “large reward vs. no money”, robust mixed ANOVA revealed a significant main effect of group (F(1,57.71)= 14.13, p<0.001), region (F(11,41.25)=2.91, p=0.01), and also an effect between group and region (F(11, 41.25)=2.61, p=0.01).

Group differences in ROI activations at 2^nd^ year follow-up between controls and risk individuals using Mann-Whitney U-tests for the contrast “large reward vs. no money” are displayed in Supplementary Table 2.

**Supplementary Table 2.** Group differences at 2^nd^ year follow-up (large reward vs. no money)

| ROI | Group 1 | Group 2 | W statistic | p-value |
| --- | --- | --- | --- | --- |
| **Left Caudate** | **controls** | **risk** | **1553** | **0.02** |
| **Right Caudate** | **controls** | **risk** | **1696** | **<0.001** |
| **Left Nucleus Accumbens** | **controls** | **risk** | **1418** | **0.02** |
| Right Nucleus Accumbens | controls | risk | 1254 | 0.26 |
| Left Putamen | controls | risk | 1440 | 0.13 |
| **Right Putamen** | **controls** | **risk** | **1559** | **0.02** |
| Left Thalamus | controls | risk | 1507 | 0.05 |
| Right Thalamus | controls | risk | 1483 | 0.05 |
| **Left dlPFC** | **controls** | **risk** | **1615** | **0.001** |
| **Right dlPFC** | **controls** | **risk** | **1591** | **0.001** |
| Left vmPFC | controls | risk | 1266 | 0.07 |
| **Right vmPFC** | **controls** | **risk** | **1688** | **<0.001** |
| Note: Group differences are calculated using Mann-Whitney U-tests. Results marked in bold represent a significant effect (p<0.05). | | | | |

**Differences in neural activation during reward anticipation: Small reward vs no money baseline**

Investigating differences in neural activation during reward anticipation at baseline for the contrast “small reward vs. no money”, robust mixed ANOVA revealed a significant main effect of region (F(11,37.49)=4.18, p<0.001), but no main effect for group or interaction effect.

Group differences in ROI activations at baseline between controls and risk individuals using Mann-Whitney U-tests for the contrast “small reward vs. no money” are displayed in Supplementary Table 3.

**Supplementary Table 3.** Group differences at baseline (small reward vs. no money)

| ROI | Group 1 | Group 2 | W statistic | p-value |
| --- | --- | --- | --- | --- |
| Left Caudate | controls | risk | 1226 | 0.26 |
| Right Caudate | controls | risk | 1199 | 0.18 |
| Left Nucleus Accumbens | controls | risk | 1112 | 0.82 |
| Right Nucleus Accumbens | controls | risk | 1006 | 0.7 |
| Left Putamen | controls | risk | 1102 | 0.35 |
| Right Putamen | controls | risk | 1093 | 0.95 |
| Left Thalamus | controls | risk | 1072 | 0.63 |
| Right Thalamus | controls | risk | 983 | 0.46 |
| Left dlPFC | controls | risk | 1085 | 0.42 |
| Right dlPFC | controls | risk | 1054 | 0.99 |
| Left vmPFC | controls | risk | 1009 | 0.84 |
| Right vmPFC | controls | risk | 1045 | 0.91 |
| Note: Group differences are calculated using Mann-Whitney U-tests. Results marked in bold represent a significant effect (p<0.05). | | | | |

**Differences in neural activation during reward anticipation: Small reward vs no money 2^nd^ year follow-up**

Investigating differences in neural activation during reward anticipation at 2^nd^ year follow-up for the contrast “small reward vs. no money”, robust mixed ANOVA did not reveal any significant effect.

Group differences in ROI activations at 2^nd^ year follow-up between controls and risk individuals using Mann-Whitney U-tests for the contrast “small reward vs. no money” are displayed in Supplementary Table 4.

**Supplementary Table 4.** Group differences at 2^nd^ year follow-up (small reward vs. no money)

| ROI | Group 1 | Group 2 | W statistic | p-value |
| --- | --- | --- | --- | --- |
| Left Caudate | controls | risk | 1435 | 0.09 |
| Right Caudate | controls | risk | 1408 | 0.09 |
| Left Nucleus Accumbens | controls | risk | 1487 | 0.07 |
| Right Nucleus Accumbens | controls | risk | 1272 | 0.21 |
| Left Putamen | controls | risk | 1366 | 0.17 |
| Right Putamen | controls | risk | 1186 | 0.23 |
| Left Thalamus | controls | risk | 1426 | 0.11 |
| Right Thalamus | controls | risk | 1291 | 0.52 |
| **Left dlPFC** | **controls** | **risk** | **1343** | **0.04** |
| Right dlPFC | controls | risk | 1191 | 0.29 |
| **Left vmPFC** | **controls** | **risk** | **1463** | **0.01** |
| Right vmPFC | controls | risk | 1129 | 0.99 |
| Note: Group differences are calculated using Mann-Whitney U-tests. Results marked in bold represent a significant effect (p<0.05). | | | | |
